# Immune Modulation in Solid Tumors: A Phase 1b Study of RO6870810 (BET Inhibitor) and Atezolizumab (PD-L1 Inhibitor)

**DOI:** 10.1101/2024.07.28.24309665

**Authors:** Daniel Marbach, Jurriaan Brouer-Visser, Laura Brennan, Sabine Wilson, Iakov I Davydov, Nicolas Staedler, José Duarte, Iris Martinez Quetglas, Eveline Nüesch, Marta Cañamero, Evelyne Chesné, George Au-Yeung, Erika Hamilton, Stephanie Lheureux, Debra L Richardson, Iben Spanggaard, Bruno Gomes, Izolda Franjkovic, Mark DeMario, Martin Kornacker, Katharina Lechner

**Affiliations:** Roche Pharma Research and Early Development, Pharmaceutical Sciences, Roche Innovation Center Basel, F. Hoffmann-La Roche Ltd, Basel, Switzerland; Roche Pharma Research and Early Development, Roche Innovation Center New York, F. Hoffmann-La Roche Ltd, New York, NY, USA; Product Development, Data Sciences, Roche Innovation Center Basel, F. Hoffmann-La Roche Ltd, Basel, Switzerland; Roche Pharma Research and Early Development, Roche Innovation Center Munich, F. Hoffmann-La Roche Ltd, Penzberg, Germany; Peter MacCallum Cancer Centre and Sir Peter MacCallum Department of Oncology, The University of Melbourne, Melbourne, Victoria, Australia; Sarah Cannon Research Institute, Nashville, TN, USA; Division of Medical Oncology and Hematology, Princess Margaret Cancer Centre, University Health Network, University of Toronto, Toronto, ON, Canada; Division of Gynecologic Oncology, Stephenson Cancer Center, University of Oklahoma Health Sciences Center, Oklahoma City, OK, United States; Department of Oncology, Rigshospitalet, Copenhagen University Hospital, Copenhagen, Denmark; Roche Pharma Research and Early Development, Oncology Early Clinical Development, Roche Innovation Center Basel, F. Hoffmann-La Roche Ltd, Basel, Switzerland

**Keywords:** bromodomain, BET inhibitor, immunotherapy, phase Ib solid tumors, TNBC, ovarian cancer

## Abstract

**Purpose:** Bromodomain and extra-terminal domain (BET) inhibitors (BETi) have demonstrated epigenetic modulation capabilities, specifically in transcriptional repression of oncogenic pathways. Preclinical assays suggest that BETi potentially attenuates the PD1/PD-L1 immune checkpoint axis, supporting its combination with immunomodulatory agents.

**Patients and Methods:** A Phase 1b clinical trial was conducted to elucidate the pharmacokinetic and pharmacodynamic profiles of the BET inhibitor RO6870810, as monotherapy and in combination with the PD-L1 antagonist atezolizumab, in patients with advanced ovarian carcinomas and triple-negative breast cancer (TNBC). Endpoints included maximum tolerated dosages, adverse event profiling, pharmacokinetic evaluations, and antitumor activity. Pharmacodynamic and immunomodulatory effects were assessed in tumor tissue (by immunohistochemistry and RNA-seq) and in peripheral blood (by flow cytometry and cytokine analysis).

**Results:** The study was terminated prematurely due to a pronounced incidence of immune-related adverse effects in patients receiving combination of RO6870810 and atezolizumab. Anti-tumor activity was limited to 2 patients (5.6%) showing partial response. Although target engagement was confirmed by established BETi pharmacodynamic markers in both blood and tumor samples, BETi failed to markedly decrease tumor PD-L1 expression and had a suppressive effect on anti-tumor immunity. Immune effector activation in tumor tissue was solely observed with the atezolizumab combination, aligning with this checkpoint inhibitor’s recognized biological effects.

**Conclusions:** The combination of BET inhibitor RO6870810 with the checkpoint inhibitor atezolizumab presents an unfavorable risk-benefit profile for ovarian cancer and TNBC (triple-negative breast cancer) patients due to the increased risk of augmented or exaggerated immune reactions, without evidence for synergistic anti-tumor effects.

**Trial registration:** ClinicalTrials.gov ID NCT03292172

## BACKGROUND

Epigenetic modifications are fundamental in guiding gene expression patterns, and alterations in these modifications are frequently associated with the onset of various malignancies(1). One prominent mechanism of epigenetic regulation is the reversible acetylation of histones, which allows for dynamic gene expression modulation in response to various stimuli. At the heart of this process is the Bromodomain and extra-terminal domain (BET) protein family, which includes BRD2, BRD3, and BRD4, and the testis-specific BRDT. Serving as epigenetic “readers”, these proteins specifically identify and bind to acetylated histones(2).

Recently, the therapeutic promise of BET protein inhibition has emerged, leading to the development of small molecule BET inhibitors (BETi), such as JQ1, which acts by binding the bromodomains of BET proteins, inhibiting their chromatin association and thereby modulating gene expression(3, 4). RO6870810 (also known as RG6146 or TEN-010), is a new non-covalent BETi. Despite its structural resemblance to JQ1, RO6870810 has superior solubility and stability, potentially offering therapeutic advantages (5).

BRD4, a primary target of RO6870810, is a universal gene transcription regulator(6). It has been linked to the upregulation of oncogenes like MYC, BCL2, CDK6, and FOSL1(7–10). Notably, BRD4 preferentially binds to super-enhancers, which are vast regulatory regions known for controlling genes necessitating high expression levels(11, 12). While the sensitivity to BETi isn’t solely dictated by super-enhancers(6, 13), genes adjacent to these regions may be linked to BRD4 inhibition.

In hematological malignancies, particularly those with MYC and BCL2 overexpression due to super-enhancer-driven transcriptional control, BETi has shown moderate success(14, 15). Accumulating evidence also suggests potential BETi susceptibility in solid tumors, like triple-negative breast cancer (TNBC) and advanced ovarian cancer which presently need effective treatments. Notably, BRD4 amplification has been documented in these cancers(16), and MYC amplification is prevalent in recurring ovarian tumors(17). Furthermore, BET proteins’ roles in immune function have potential utility in cancer therapy. While early research highlighted JQ1’s ability to suppress immune regulators in various tumor models(18–20), newer preclinical studies showcase BETi’s diverse impacts on immune cell subtypes and activation.

The safety and efficacy of the BET inhibitor RO6870810 combined with venetoclax and rituximab was previously investigated for the treatment of relapsed or refractory diffuse large B-cell lymphoma (DLBCL) (20). In this phase 1b trial involving 39 patients, the combination therapy showed tolerability with manageable toxicities. Dose-limiting toxicities included neutropenia, diarrhea, and hyperbilirubinemia. The maximum tolerated dose (MTD) for the combination of RO6870810 and venetoclax was established at 0.65 mg/kg for RO6870810 and 600 mg for venetoclax. For the triple combination of RO6870810, venetoclax, and rituximab, the MTDs were 0.45 mg/kg, 600 mg, and 375 mg/m², respectively. The combination showed promising anti-tumor activity with an overall response rate of 38.5% and complete responses in 20.5% of patients.

Another phase 1b trial was conducted to determine the maximum tolerated dose (MTD) and optimal biological dose (OBD) of RO6870810 monotherapy in patients with advanced multiple myeloma (21). Though pharmacodynamic results indicated the on-target effects of RO6870810, clinical responses were infrequent and, when present, transient. These findings align with the preliminary activity noted for RO6870810 in an earlier first-in-human dose-escalation study. There, objective response rates (ORRs) stood at 25% (2/8) for nuclear protein of the testis carcinoma (NUT carcinoma), 2% (1/47) for other solid tumors, and 11% (2/19) for diffuse large B-cell lymphoma (DLBCL)(21).

In a study by Roboz G.J. et al.(22), 32 patients with relapsed/refractory acute myeloid leukemia and hypomethylating agent–refractory myelodysplastic syndrome were treated with RO6870810 monotherapy(22). Significant reductions in circulating CD11b+ cells, a known pharmacodynamic marker of BET inhibition, were observed at RO6870810 concentrations exceeding 120 ng/mL. Most side effects were mild, and there were no treatment-related fatalities. Although some patients showed signs of stabilization or remission, the development of RO6870810 as a standalone therapy was discontinued due to its limited efficacy.

The ability of BETi to inhibit the PD-1/PD-L1 immune checkpoint pathway and bolster anti-tumor immunity suggests that combining it with a checkpoint inhibitor could yield improved clinical outcomes(23). Supporting this notion, preclinical studies using a combination of BETi with anti-PD-1 or anti-PD-L1 antibodies have showcased synergistic anti-tumor effects in mouse models of lymphoma(19), melanoma(24), and non-small cell lung cancer(25). Yet, clinical evidence from such combination therapies remains unreported(23, 26).

In this study, we present findings from a phase 1b clinical trial involving TNBC and ovarian cancer patients. These patients received treatment with the BETi RO6870810 as a monotherapy or in combination with atezolizumab (Tecentriq), a humanized IgG1 monoclonal antibody targeting PD-L1. Notably, atezolizumab has secured approval for treating PD-L1 positive metastatic TNBC(27). Our study examines the potential anti-tumor immune activation facilitated by both RO6870810 monotherapy and its combination with atezolizumab. We offer a detailed biomarker analysis, highlighting transcriptional alterations and immune modulation in both tumor tissue and peripheral blood. This is the first study to explore the effects of combining BET inhibition with PD-L1 blockade to enhance therapeutic efficacy by targeting both the epigenetic regulation pathways and immune checkpoint pathways simultaneously.

## METHODS

### Study design

We conducted a phase 1b, open-label, non-randomized trial on patients ≥18 years with TNBC and advanced ovarian cancer. We explored two treatment strategies: (1) immediate combination of RO6870810 administered subcutaneously, with intravenous atezolizumab (concomitant regimen, Fig. 1a), and (2) an initial 21-day single-agent, subcutaneous RO6870810 treatment, followed by its combination with intravenous atezolizumab (sequential regimen, Fig. 1b). The dose-escalation followed a classic 3+3 design with initially planned doses of 0.30 mg/kg, 0.45 mg/kg, and 0.65 mg/kg. The study had four groups. Groups 1 and 2 focused on dose escalation for the concomitant and sequential treatments, respectively. Patients in group 1 received a starting-dose of 0.30 mg/kg for 14 days administered subcutaneously on a 3-week schedule. Once a cohort in group 1 was completed and deemed safe, group 2 began the 21-day run-in period, during which RO6870810 monotherapy was administered to a minimum of 3 participants. Participants enrolled in group 2 initially received RO6870810 as monotherapy during the first 14 days of a 21-day run-in period, starting at a dose of 0.30 mg/kg. Patients in the same dose level were treated simultaneously. Following the run-in period, participants continued to receive RO6870810 at the same dose in combination with 1200 mg atezolizumab in 21-day cycles. In the expansion phase, Cohorts 3 and 4 further investigated the concomitant regime for TNBC and ovarian cancer patients, using the optimal dose determined in Cohort 1.

**Figure 1.**
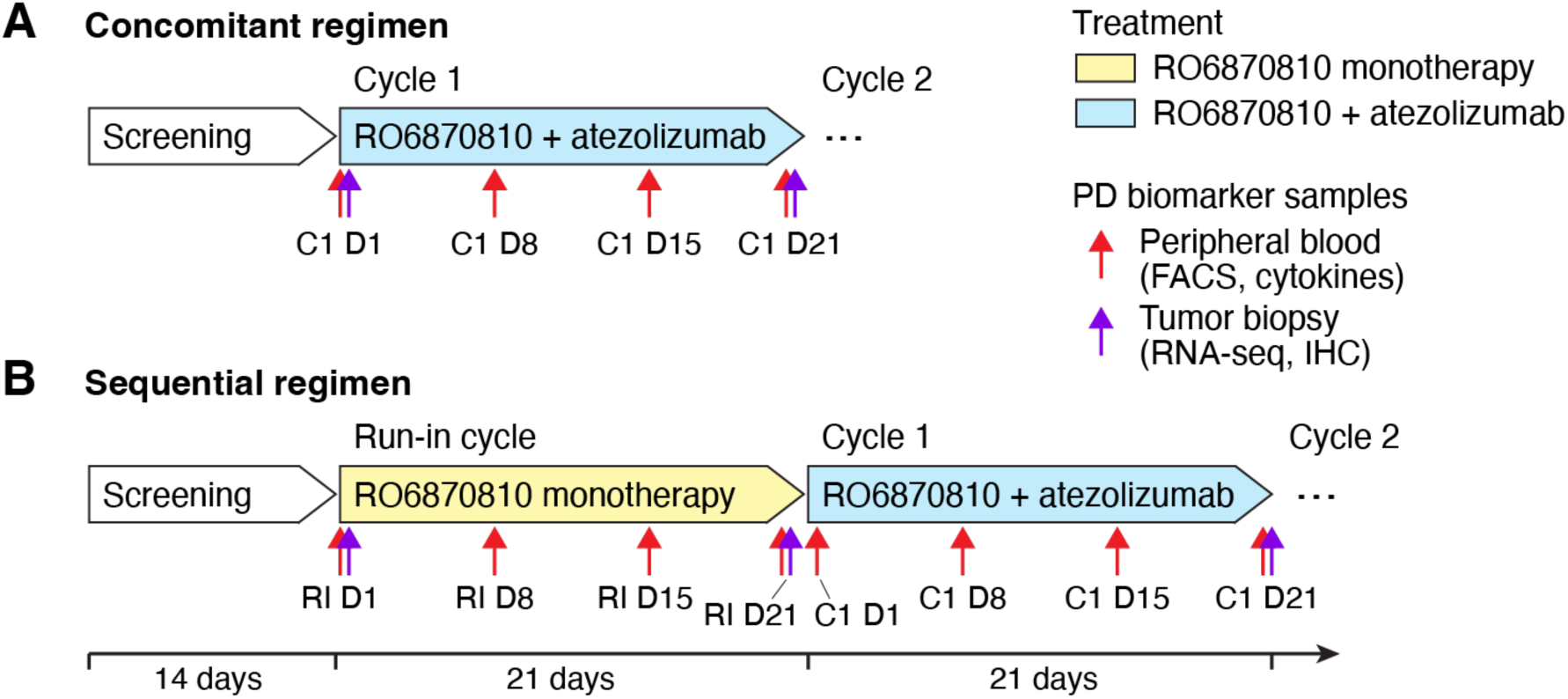
Schematic Overview of Study Treatment Regimens and Pharmacodynamic Biomarker Collection. **A.** The concomitant regimen involved patients receiving a combination of RO6870810 and atezolizumab from initiation. Tumor biopsies for RNA-sequencing and immunohistochemistry (IHC) were taken at baseline (Cycle 1 Day 1 [C1 D1]) and post-first cycle (Cycle 1 Day 21 [C1 D21]), indicated by purple arrows. Peripheral blood samples for flow cytometry and cytokine profiling, shown by red arrows, were collected on days 1, 8, 15, and 21. This regimen was applied to patients in the dose escalation and both expansion cohorts. **B.** To evaluate the impact of RO6870810 as a single agent, an alternative group followed a sequential regimen, starting with RO6870810 alone in a run-in cycle before transitioning to combined treatment with atezolizumab. Tumor biopsies were performed at the run-in start (Run-in Day 1 [RI D1]), post-run-in cycle (Run-In Day 21 [RI D21]), and after the initial cycle of combination therapy (C1 D21). Peripheral blood sampling occurred on the same days during the run-in and the first combination treatment cycle, facilitating a comprehensive analysis of treatment-induced changes.

The study primarily aimed to ascertain the maximum tolerated dose (MTD) or maximum administered dose (MDA) of RO6870810 both as a standalone treatment and in combination with atezolizumab, by monitoring dose-limiting side effects and ongoing safety. The expansion groups enabled us to gauge the early clinical efficacy of RO6870810 when paired with atezolizumab. Additionally, understanding the immune modulation profiles of RO6870810, both as monotherapy and when combined with PD-L1 inhibition, was a goal for this study.

Objective responses were assessed by investigators according to RECIST v1.1 and Immune Modified RECIST criteria. The grading of all adverse events (AEs) was based on the National Cancer Institute Common Toxicity Criteria for Adverse Events (NCI CTCAE) version 4.03.

The study’s methodology, eligibility criteria, dosing schedules, and safety protocols are detailed in the Supplementary Methods. Further information is accessible on ClinicalTrials.gov under trial ID NCT03292172 or via this direct link: https://clinicaltrials.gov/study/NCT03292172.

### Sample collection and analysis

Blood samples were collected at specified intervals for biomarker analysis. Flow cytometry was conducted at Covance Central Laboratory using established protocols. Cytokine levels were measured using the ELLA method by Microcoat Biotechnologie. Tumor biopsies were processed for immunohistochemistry and RNA-seq to study gene expression and pathway activity. The detailed methods, including sample preparation, analytical procedures, and statistical analyses, are provided in the supplementary methods section.

## RESULTS

### Patient demographics and key clinical data

#### Dosing of RO6870810

The dosing rationale was based on pharmacokinetic profile and tolerability of RO6870810 observed in patients with NUT carcinoma, other solid tumors, and DLBCL(21). In this study, RO6870810 demonstrated overall tolerability across different indications except for a single dose-limiting toxicity (DLT) of grade 3 cholestatic hepatitis observed in a patient with prostate cancer at 0.45 mg/kg on a 28-day schedule. This led to the expansion of the cohort without additional DLTs and dose escalation to 0.65 mg/kg. Although no DLTs were reported at this level during cycle 1, treatment discontinuations due to fatigue in cycle 2 prompted the exploration of a 14 of 21 days schedule. This 0.65 mg/kg dose was identified as the recommended phase 2 dose for solid tumors. Similarly, in the study by Dickinson et al.(15), the maximum tolerated dose (MTD) for the combination of RO6870810 and venetoclax was established at 0.65 mg/kg for RO6870810 and 600 mg for venetoclax. For the triple combination of RO6870810, venetoclax, and rituximab, the MTDs were determined to be 0.45 mg/kg for RO6870810, 600 mg for venetoclax, and 375 mg/m² for rituximab.

Based on the safety profile and pharmacodynamic (PD) effects observed, a starting dose of 0.3 mg/kg for 14 days on a 3-week schedule was selected as appropriate for the initial dose cohort of both groups. This dosage was anticipated to provide significant target PD effects while maintaining a tolerable safety profile. This strategy aimed to optimize the therapeutic potential of RO6870810 in combination with atezolizumab for the patient population in this study.

#### Participants

Thirty-six (36) patients with metastatic advanced ovarian cancer (n= 29) or triple negative breast cancer (n= 7) were included and received at least one dose of study drug in this open-label, dose finding and expansion phase 1 study. The total of 36 safety evaluable patients were enrolled in Denmark (8 patients), Canada (10 patients), the US (15 patients), and Australia (3 patients). Details of the groups and cohorts and their dosages are provided in Table 1.

**Table 1.**
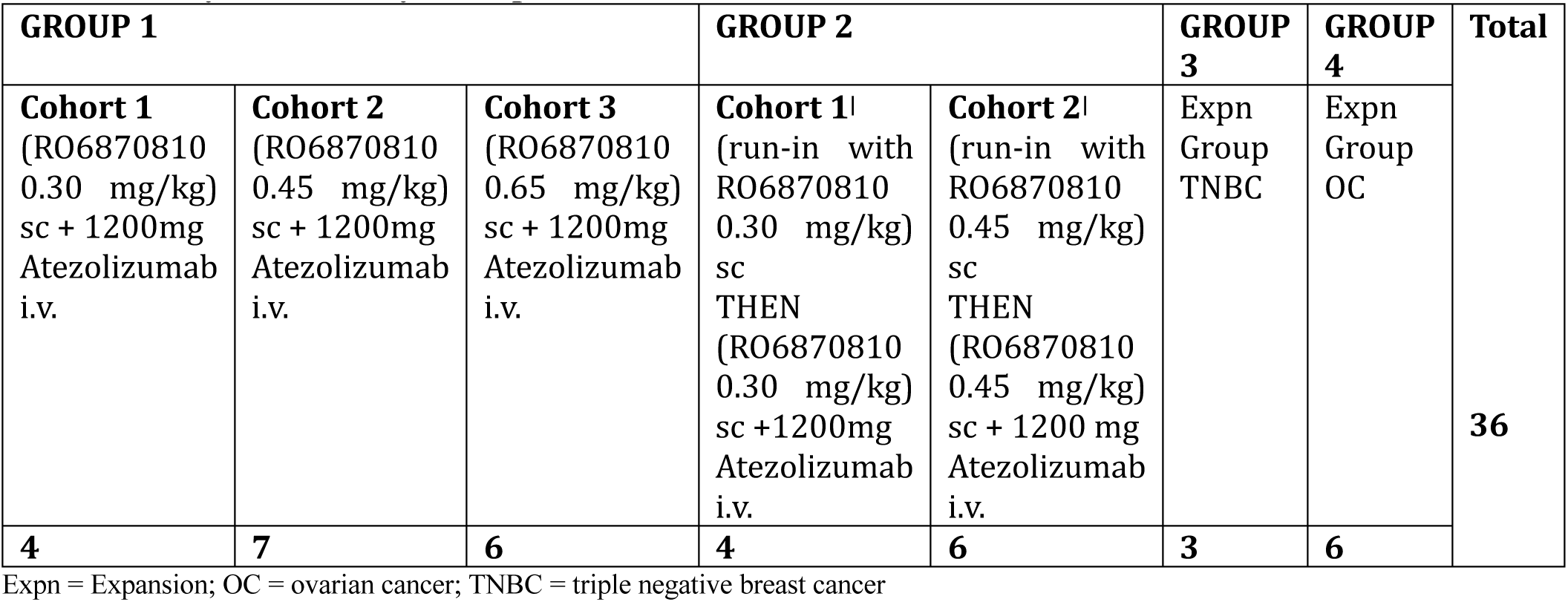
Study Patients, by Group.

Twenty-seven patients were included in the dose escalation part (groups 1 and 2) and 9 patients were treated in the expansion phase at the recommended phase 2 dose of 0.45mg/kg. The median age of all enrolled female patients was 53 years (range: 34-72 years) with 22 patients (61.1%) showing an ECOG score of 1 and 14 patients (38.9%) an ECOG score of 0. All 36 enrolled patients discontinued the study treatment; the primary reasons for treatment discontinuation were progressive disease (21 patients [58.3%]) and AEs (8 patients [22.2%]). Of the 36 patients enrolled, 29 patients discontinued and 7 patients completed the study. The primary reasons for study discontinuation were death (12 patients [33.3%]), followed by a reason of “other” (7 patients [19.4%]). Further reasons of study discontinuation were withdrawal by the patient (6 patients [16.7%], progressive disease and study terminated by Sponsor (2 patients [5.6%]).

#### Safety

The study was terminated prematurely because of frequency and severity of adverse events (AEs) and an unfavorable risk-benefit profile of the combination of RO6870810 and Atezolizumab. All participants (100%, 36/36) experienced at least one AE, with 97.2% (35/36) reporting treatment-related AEs. A total of 473 AEs were documented. Discontinuation due to AEs affected 22.2% (8/36) of patients.

Grade ≥3 AEs were reported in 63.9% (23/36) of patients, with serious adverse events (SAEs) occurring in 58.3% (21/36). Of these, treatment-related Grade ≥3 AEs were observed in 41.7% (15/36) of patients, and treatment-related SAEs in 33.3% (12/36). One dose-limiting toxicity (DLT) was identified at dose level 3, attributed to a Grade 3 systemic immune activation event in one patient from Group 1, Cohort 3, at a dosage of 0.65 mg/kg in combination with atezolizumab. This event, deemed related to the study treatment, led to the discontinuation of treatment for this patient.

Among the 21 patients (58.3%) who experienced SAEs, a total of 35 SAEs were reported. SAEs occurring in ≥5% of patients included systemic immune activation (4 patients [11.1%]), small intestinal obstruction (3 patients [8.3%]), abdominal pain, chest pain, fatigue, and pyrexia (each reported by 2 patients [5.6%]).

The system organ classes (SOCs) in which AEs were experienced by ≥50% patients were: General disorders and administration site conditions (35 patients [97.2%]), gastrointestinal disorder (29 patients [80.6%]), metabolism and nutrition disorder (23 patients [63.9%]), and respiratory, thoracic and mediastinal disorders (19 patients [52.8%] each). The most frequently reported AEs, affecting at least 30% of participants, included fatigue and injection site reactions (66.7%, 24/36 for each Preferred Term [PT]), diarrhea (50.0%, 18/36), nausea (44.4%, 16/36), decreased appetite (41.7%, 15/36), pyrexia (36.1%, 13/36), and vomiting (30.6%, 11/36). Adverse events related to the study treatment and reported by at least 30% of the patients, were injection site reactions (66.7%, 24/36), fatigue (52.8%, 19/36), diarrhea (41.7%, 15/36), decreased appetite (36.1%, 13/36), and nausea (33.3%, 12/36).

The study recorded 15 deaths (41.7%), with nine deaths due to progressive disease and six deaths (16.7%) reported during long-term follow-up where the cause of death was unknown. None of the deaths were treatment-related.

Although there were laboratory abnormalities in both hematological (high and low) and clinical chemistry (high and low) parameters, none of these were considered clinically significant. No clinically meaningful differences from baseline were noted in the vital signs.

#### Efficacy

Response was measured according to RECIST overall response. Out of 31 evaluable patients, two patients exhibited a partial response (PR), fifteen patients demonstrated stable disease (SD), and fourteen patients were classified with progressive disease (PD) as their best objective response (Figure 2). Further breakdown and detailed analysis of patient responses across different groups and cohorts are documented in Table 2.

**Figure 2:**
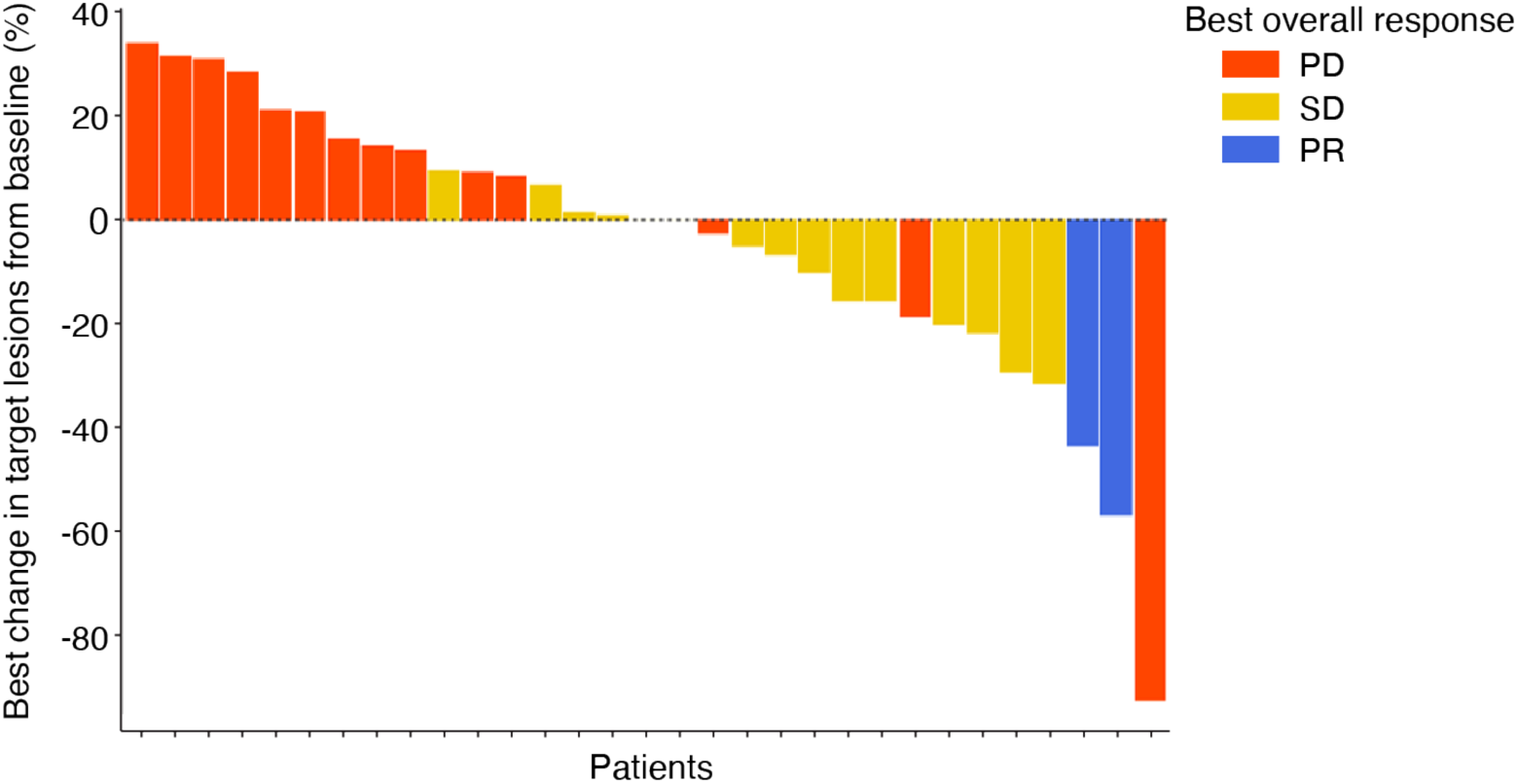
Changes in Target Lesion Size and Best Overall Response. Each bar represents the response of an individual patient, measured according to RECIST overall response criteria. The y-axis corresponds to the maximum percentage change from baseline in sum of longest diameters (SLD) in target lesions. Colors indicate the best overall response. Out of 36 patients, 31 were evaluable for clinical response. Two patients who exhibited a decrease in target lesion size were still classified as having progressive disease due to progression in non-target lesions or the appearance of new lesions.

**Table 2.**
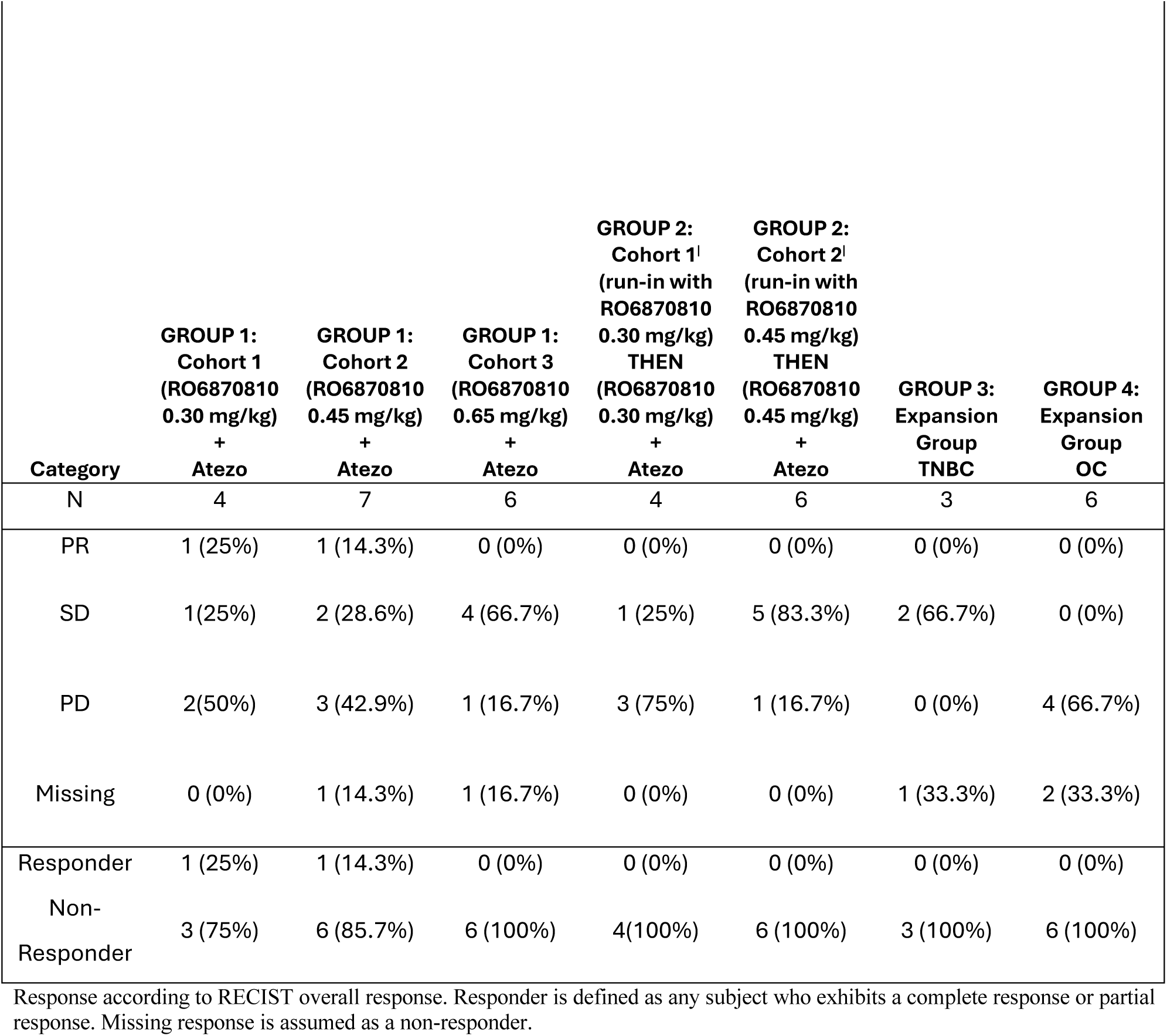

| Category | GROUP 1:<br>Cohort 1<br>(RO6870810<br>0.30 mg/kg)<br>+<br>Atezo | GROUP 1:<br>Cohort 2<br>(RO6870810<br>0.45 mg/kg)<br>+<br>Atezo | GROUP 1:<br>Cohort 3<br>(RO6870810<br>0.65 mg/kg)<br>+<br>Atezo | GROUP 2:<br>Cohort 1<sup>1</sup><br>(run-in with<br>RO6870810<br>0.30 mg/kg)<br>THEN<br>(RO6870810<br>0.30 mg/kg)<br>+<br>Atezo | GROUP 2:<br>Cohort 2<sup>1</sup><br>(run-in with<br>RO6870810<br>0.45 mg/kg)<br>THEN<br>(RO6870810<br>0.45 mg/kg)<br>+<br>Atezo | GROUP 3:<br>Expansion<br>Group<br>TNBC | GROUP 4:<br>Expansion<br>Group<br>OC |
| --- | --- | --- | --- | --- | --- | --- | --- |
| N | 4 | 7 | 6 | 4 | 6 | 3 | 6 |
| PR | 1 (25%) | 1 (14.3%) | 0 (0%) | 0 (0%) | 0 (0%) | 0 (0%) | 0 (0%) |
| SD | 1(25%) | 2 (28.6%) | 4 (66.7%) | 1 (25%) | 5 (83.3%) | 2 (66.7%) | 0 (0%) |
| PD | 2(50%) | 3 (42.9%) | 1 (16.7%) | 3 (75%) | 1 (16.7%) | 0 (0%) | 4 (66.7%) |
| Missing | 0 (0%) | 1 (14.3%) | 1 (16.7%) | 0 (0%) | 0 (0%) | 1 (33.3%) | 2 (33.3%) |
| Responder | 1 (25%) | 1 (14.3%) | 0 (0%) | 0 (0%) | 0 (0%) | 0 (0%) | 0 (0%) |
| Non-Responder | 3 (75%) | 6 (85.7%) | 6 (100%) | 4(100%) | 6 (100%) | 3 (100%) | 6 (100%) |

Response according to RECIST overall response. Responder is defined as any subject who exhibits a complete response or partial response. Missing response is assumed as a non-responder.

The two partial responses were observed in Group 1, Cohort 1, which received a dosage of 0.3 mg/kg concurrently, and in Group 1, Cohort 2, with a 0.45 mg/kg concurrent dosage. Five patients were excluded from the clinical response evaluation due to the absence of post-baseline response data and were therefore categorized as having progressive disease.

### Pharmacodynamic effects for BETi biomarkers

Pharmacodynamic (PD) biomarkers for RO6870810 were evaluated in peripheral blood and tumor tissue. BET inhibitors are known to target peripheral blood monocytes(28), which are critical determinants of cancer-associated inflammation. A previous study with RO6870810 has suggested that circulating monocyte levels in peripheral blood can be used as a potential biomarker for the pharmacodynamic effects(29). Figure 3A presents the results of circulating monocytes following concurrent or sequential administration of RO6870810 and atezolizumab. We observed a significant decrease in CD14+/CD11b+ monocytes after the initial treatment cycle, with the lowest counts recorded between days 8 and 14 post-treatment. These levels then recovered by day 21.

**Figure 3.**
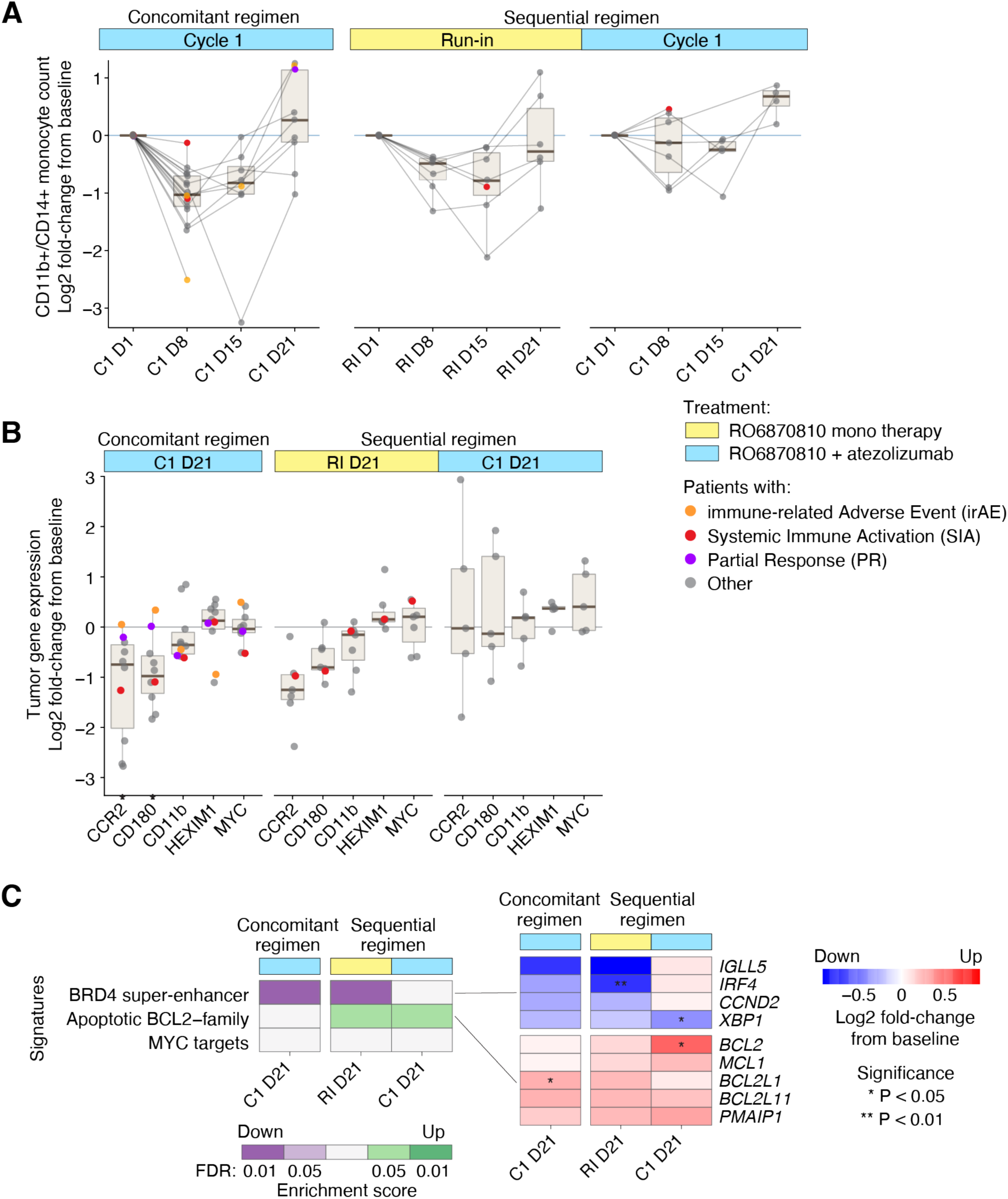
Pharmacodynamic Responses of BET Inhibitor Biomarkers in Peripheral Blood and Tumor Tissue. **A.** Quantification of CD14+/CD11b+ monocyte populations in peripheral blood, illustrating changes from baseline (expressed as log2 fold-change from cycle onset) for individual patients (denoted as points), with longitudinal data from the same individual linked. Patients lacking baseline or sequential samples are excluded. Color highlights patients with partial response (purple), immune-mediated adverse events (orange), or systemic immune activation (red). Refer to Fig. 1 for time point definitions. Boxplots depict median (center line), quartiles (box limits), and variability (whiskers extend to 1.5x interquartile range). **B.** Tumor expression levels of established BETi target genes, as determined by RNA-seq, indicating gene expression modifications (log2 fold-change) from the screening (pre-treatment) sample. Exclusions apply for participants without screening or on-treatment samples. The same color coding as in Panel A is used. **C.** Gene signature enrichment analysis reflecting BETi downstream effects, with heatmaps showcasing signature scores and gene expression alterations. Green and purple denote significantly up- or down-regulated signatures, respectively, with red and blue highlighting individual gene expression shifts within significant signatures. Asterisks indicate statistical significance. Time points align with those in Panel B.

We further investigated the expression of genes affected by BET inhibitors (BETi) within the tumor tissue using RNA sequencing (RNA-seq). The genes CD180, CCR2, MYC and HEXIM1 are previously reported pharmacodynamic markers of BETi in different settings(26). On day 21, significant reductions in the levels of CCR2 and CD180 were confirmed under both the concurrent regimen and the monotherapy initiation with RO6870810, while MYC and HEXIM1 were not significantly affected (Figure 3B). The treatment also led to the downregulation of the BRD4 super enhancer, alongside specific changes in the expression of apoptotic and BCL2 family genes (Figure 3C). Notably, BCL2 and BCL2L1 were upregulated, whereas IGLL5 and IRF4 were downregulated. These gene expression changes, particularly within the context of apoptosis and lymphocyte regulation, underscore the potential mechanisms through which RO6870810 exerts its anti-tumor effects.

We also examined the changes in cellular subsets and soluble biomarkers within peripheral blood as assessed by flow cytometry and cytokine profiling. Besides the decrease in CD14+/CD11b+ monocytes discussed above, no notable changes were observed for the run-in cycle with RO6870810 alone. In contrast, early phases of the combination therapy with atezolizumab were characterized by a transient reduction in circulating immune cells, including CD4+ and CD8+ cells, CD16+CD56+ NK cells, CD19+ B cells, and CD14+/CD11b+ monocytes (Figure 4A). The transient drop in circulating immune cells, potentially due to margination and extravasation, has been previously described for other immunotherapeutic modalities involving T cell activation(30, 31). Following this initial reduction in circulating immune cells, there was an expansion of specific cell types, particularly CD16+CD56+ NK cells and CD8+ T cells, but not CD4+ T cells (Figure 4A). Consequently, the ratio of CD4+ to CD8+ T cells shifted towards a higher proportion of cytotoxic cells in the later phase of the combination therapy (Figure 4B).

**Figure 4.**
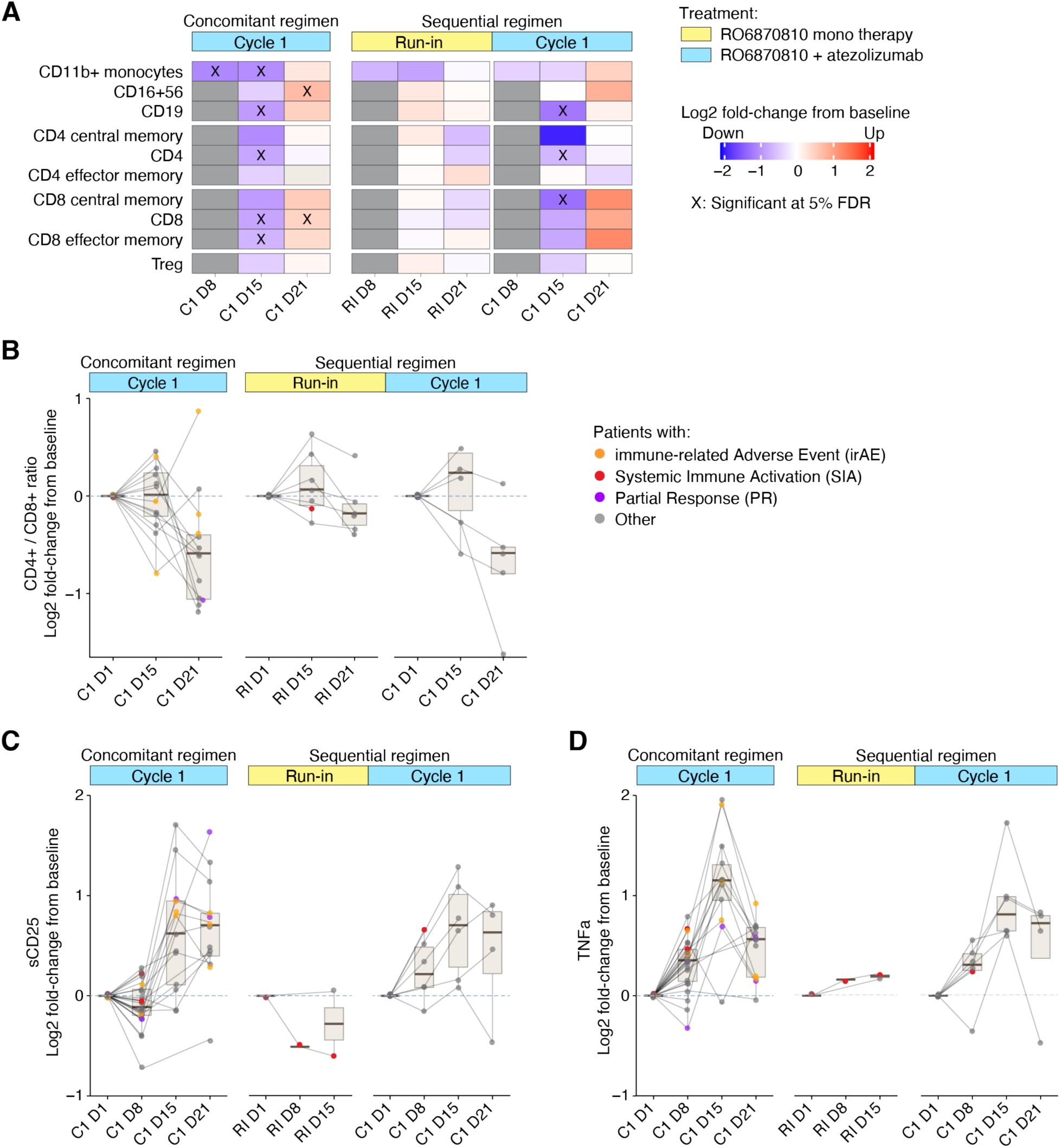
Assessment of Immune Modulation by Flow Cytometry and Cytokine Analyses. **A.** The variation in immune cell populations within peripheral blood, as determined by flow cytometry. Color depicts the log2 fold-change from baseline at each defined time point (refer to Fig. 1 for time points). Red indicates an increase, blue a decrease in cell population frequency, with significant alterations marked by an ‘X’ (FDR corrected p-value < 0.05). **B.** Change from baseline in the CD4+/CD8+ cell ratio in peripheral blood, indicating shifts towards either T helper cells (positive values) or cytotoxic cells (negative values). Continuous lines connect sequential time point samples from individual patients, highlighting specific cases of interest in color. Boxplots aggregate data at each time point. **C, D.** Changes in soluble CD25 (sCD25) and TNFα levels from baseline in peripheral blood. The visualization follows the format of Panel B.

In the combination therapy with atezolizumab, the concentration of sCD25, a soluble form of the IL-2 receptor alpha chain, showed a marked increase on day 15 post-treatment initiation, with levels remaining elevated through day 21 (Figure 4C). This elevation in sCD25 is indicative of T cell activation, suggesting enhanced immune activation potentially conducive to antitumor activity. Similarly, TNFα, a critical cytokine in inflammation and immune regulation, exhibited a marked increase on-treatment with a peak at day 15 (Figure 4D). These effects were not observed during the run-in cycle with RO6870810 alone, suggesting that the immune-stimulating effects in the combination therapy are driven by atezolizumab.

We subsequently examined tumor tissue by RNA-seq in order to explore immune gene and signature expression changes (Figure 5). Consistent with the established mechanism of action of the PD-L1 inhibitor atezolizumab, we confirm up-regulation of immune effector gene signatures in tumor tissues under the combination therapy, including signatures associated with CD8+ T cell effector functions and antigen processing machinery. In sharp contrast, the same immune effector signatures were down-regulated in patients treated with BETi alone (Figure 5A).

**Figure 5.**
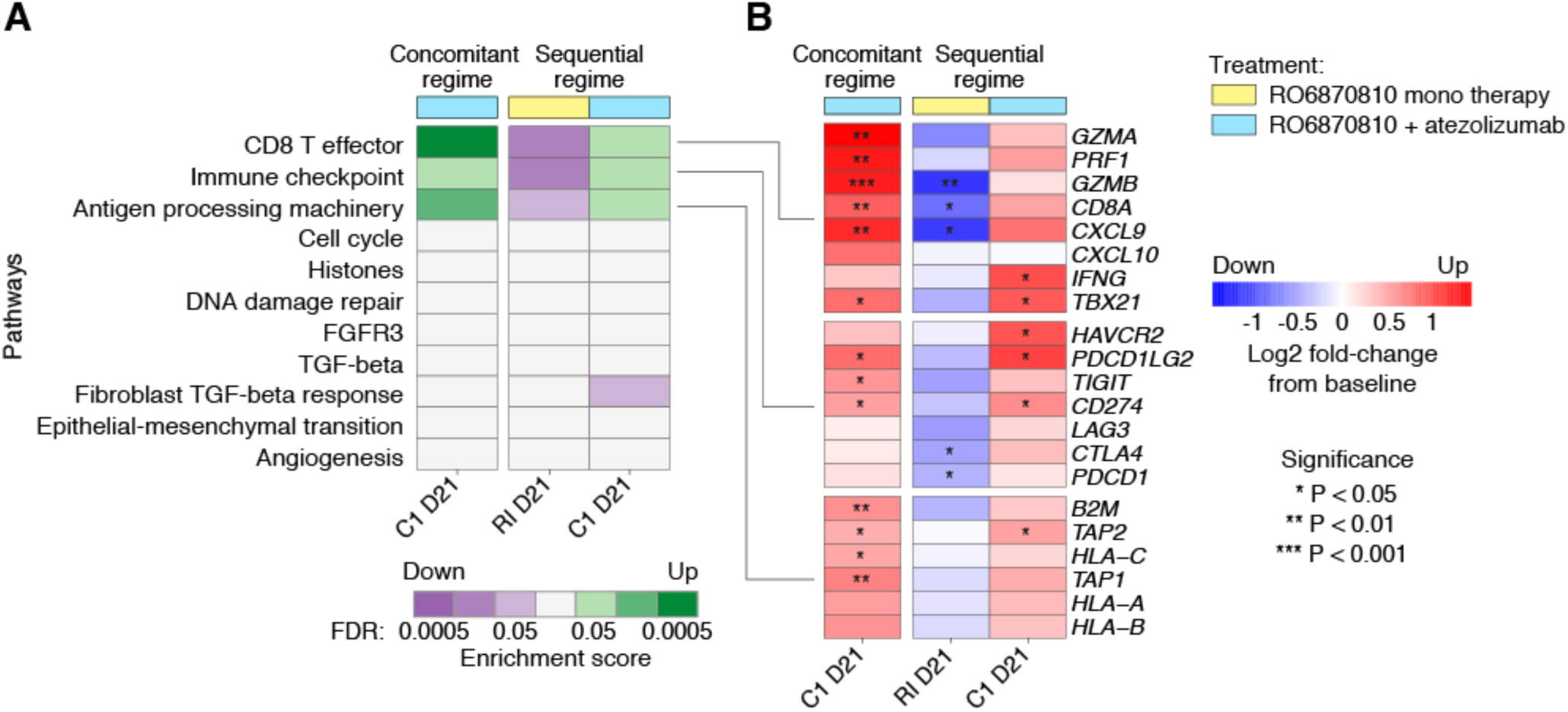
Differential Impact of BET inhibitor Monotherapy and Atezolizumab Combination Therapy on Immune Effector Pathways. Heatmaps illustrate the contrasting effects of atezolizumab combination therapy and BET inhibitor monotherapy on immune effector pathways within tumor tissues, based on RNA sequencing data. **A.** Enrichment scores for key immune pathways(34). Green indicates significant upregulation in combination therapy, suggesting enhanced immune activity. Purple marks downregulation in BETi monotherapy, implying reduced immune response. **B.** Gene expression changes related to CD8 T effector, immune checkpoint, and antigen processing machinery pathways are highlighted. Red represents upregulated genes, reflecting pathway activation, while blue indicates downregulated genes, signifying pathway suppression. Significant changes are marked with asterisk.

These observations were confirmed at the level of individual genes within those signatures (Figure 5B). We found marked increases in gene expression related to inflammation within the tumor microenvironment. For example, genes related to T cell activation and infiltration, immune surveillance, cytokine signaling, cell-mediated cytotoxicity and the IFN-γ response exhibited significant up-regulation at day 21 under the combination therapy with both treatment regimens, consistent with the induction of a robust antitumor immune environment. In contrast, a suppression of these genes and pathways was observed during the BETi monotherapy run-in, aligning with a more immunosuppressive tumor microenvironment (Figure 5B).

We further validated these findings using immunohistochemistry (IHC) data from tumor tissue samples. Contrary to pre-clinical expectations that BET inhibition would suppress PD-L1 expression and thereby enhance anti-tumor immunity(19, 20, 32, 33) treatment with RO6870810 did not reduce PD-L1 expression during the monotherapy run-in phase. Additionally, RO6870810 failed to prevent the likely IFN-γ-induced upregulation of PD-L1 when combined with atezolizumab (Supplementary Figure 1).

## DISCUSSION

This is the first study to clinically evaluate the combination of BET inhibition and immune checkpoint inhibition. Patients with advanced metastatic ovarian cancer and triple-negative breast cancer were treated with the BET inhibitor RO6870810 and the PD-L1 inhibitor atezolizumab following two alternative regimens, with or without an RO6870810 monotherapy run-in phase. Despite the promising preclinical evidence suggesting potential synergistic effects of combining BET inhibitors with checkpoint inhibitors, our phase 1b study highlights significant challenges and limitations associated with this therapeutic strategy.

Although each agent has a manageable safety profile when used alone, the combination of RO6870810 and atezolizumab led to pronounced immune-related adverse events (irAEs), necessitating premature study termination. The majority of patients experienced treatment-related adverse events, with a substantial proportion encountering severe (Grade ≥3) adverse events and serious adverse events (SAEs). Notably, systemic immune activation (SIA) was a prominent SAE, underscoring the potential for heightened immune responses when combining these agents. These findings align with the known immune-stimulatory effects of checkpoint inhibitors but suggest that the addition of BET inhibition may exacerbate these responses, leading to an unfavorable risk-benefit profile.

Pharmacodynamic analyses confirmed target engagement by RO6870810, as evidenced by changes in established BETi biomarkers in both peripheral blood and tumor tissue. However, contrary to preclinical expectations, RO6870810 monotherapy did not significantly decrease tumor PD-L1 expression and appeared to suppress anti-tumor immunity within the tumor microenvironment (TME). This immunosuppressive effect was only reversed when RO6870810 was combined with atezolizumab, which induced immune effector activation in the TME. This highlights the pivotal role of atezolizumab in stimulating anti-tumor immunity, consistent with its known mechanism of action as a PD-L1 inhibitor.

The combination therapy also induced systemic immune effects, evidenced by transient reductions in circulating immune cells followed by their expansion, and increased levels of soluble immune activation markers such as sCD25 and TNFα. These systemic changes suggest that while the combination can activate the immune system, it may also predispose patients to severe irAEs.

The observed changes in both circulating immune cells and soluble factors, following concomitant and sequential administration of the treatments, but not with the monotherapy run-in phase using RO6870810 alone, underscore the critical role of atezolizumab in eliciting the potential antitumor immune response. Atezolizumab, by enhancing immune activation and possibly improving the recognition and elimination of tumor cells, emerges as the primary driver behind the immune modulatory effects observed, rather than RO6870810.

The anti-tumor activity observed in this study was limited, with only two patients (5.6%) achieving partial responses. This modest efficacy, coupled with the high incidence of severe irAEs, further supports the conclusion that the combination of RO6870810 and atezolizumab does not provide a favorable therapeutic benefit for patients with advanced ovarian carcinomas and TNBC. The lack of significant tumor PD-L1 modulation by RO6870810 and the observed immunosuppressive effects during monotherapy suggest that BET inhibition may not enhance the efficacy of checkpoint inhibitors in these cancer types.

Our study underscores the complexity of combining epigenetic modulators with immunotherapies. The anticipated synergistic stimulation of anti-tumor immunity from combining BET inhibitors with checkpoint blockade, as suggested by preclinical models, could not be confirmed in our clinical study. This discrepancy underscores the significant challenges in translating preclinical findings to clinical settings. The observed immunosuppressive effects of BETi monotherapy within the TME suggest that we need better models and a deeper understanding of the context-dependent effects of these agents on immune modulation.

## CONCLUSIONS

The combination of RO6870810 and atezolizumab demonstrated some immune activation; however, the associated severe irAEs and limited anti-tumor efficacy indicate that this therapeutic approach is not viable for patients with advanced ovarian carcinomas and TNBC. These findings highlight the importance of careful evaluation of combination strategies in clinical trials and the need for continued exploration of novel therapeutic approaches to improve outcomes for patients with these challenging malignancies.

## Data Availability

Qualified researchers may request access to individual participant-level data through the clinical study data request platform (https://vivli.org/ourmember/roche/). Further details on F. Hoffmann-La Roche Ltd's criteria for eligible studies are available at https://vivli.org/members/ourmembers/. For further details on F. Hoffmann-La Roche Ltd's Global Policy on the Sharing of Clinical Information and the procedure to request access to related clinical study documents, see https://www.roche.com/innovation/process/clinical-trials/data-sharing

## DECLARATIONS

### Ethics approval and consent to participate

This study was approved by each center’s ethics committee or institutional review board, and the study was conducted in accordance with the principles of the Declaration of Helsinki and Good Clinical Practice guidelines. All participants provided written informed consent. List of independent Ethics Committees/Institutional Review Boards with dates of approval: (1) University Health Network Research Ethics Board, 700 Bay Street, 17th Floor, Suite 1700, M5G 1Z6, Toronto, Ontario, CANADA (Approval: 12-Oct-2017); (2) Dana Farber Cancer Institute/Dana-Farber/Harvard Cancer center, 450 Brookline Ave, OS-200, Boston, MA, 02215, UNITED STATES (Approval: 07-Nov-2017); (3) Western Institutional Review Board, 1019 39th Avenue SE, Ste 120, Puyallup, WA, 98374, UNITED STATES (Approval: 18-Oct-2017); (4) Peter MacCallum Cancer Centre Ethics Committee, 305 Grattan Street, 3000, Melbourne, Victoria, AUSTRALIA (Approval: 01-Aug-2018); (5) IntegReview Ethical Review Board, 3001 S. Lamar Blvd., Suite 210, Austin, TX, 78704, UNITED STATES (Approval: 03-Sept-2018).

### Consent for publication

All of the authors have provided consent for publication.

### Availability of data and materials

Qualified researchers may request access to individual participant–level data through the clinical study data request platform (https://vivli.org/ourmember/roche/). Further details on F. Hoffmann-La Roche Ltd’s criteria for eligible studies are available at https://vivli.org/members/ourmembers/. For further details on F. Hoffmann-La Roche Ltd’s Global Policy on the Sharing of Clinical Information and the procedure to request access to related clinical study documents, see https://www.roche.com/innovation/process/clinical-trials/data-sharing.

### Authors’ contributions

GAY, EH, SL, DR, GS and IS served as site investigators for the trial. The remaining authors made substantial contributions to the study’s design and methodology, clinical and laboratory assessments, data acquisition, and analysis. They were also involved in developing the statistical approaches, conducting biomarker analyses, and interpreting the results of the trial. Acknowledgements: We thank the patients and their families for participating in the clinical trial. Editorial assistance was provided by Dr. A. Raja Choudhury.

