## Supplementary Information for "Immune Modulation in Solid Tumors: A Phase 1b Study of RO6870810 (BET Inhibitor) and Atezolizumab (PD-L1 Inhibitor)"

Supplementary methods, figures and tables of manuscript: Marbach D, Brouwer-Visser J et al., Immune Modulation in Solid Tumors: A Phase 1b Study of RO6870810 (BET Inhibitor) and Atezolizumab (PD-L1 Inhibitor), 2024,

### Supplementary Methods

#### Patients

Patients aged 18 years and older diagnosed with either ovarian cancer or triple-negative breast cancer (TNBC) were categorized into groups 1 and 2. In Groups 1 and 2, patients who had received prior treatment with CD137 agonists or immune checkpoint blockade therapies, including anti-cytotoxic T-lymphocyte-associated protein 4 (CTLA-4), anti-Programmed cell death-1 (PD-1), and anti- Programmed death-ligand 1 (PD-L1) therapeutic antibodies, could be enrolled, after a minimum of 5 months from the last dose and provided they had no history of severe immune-related adverse effects from CD137 agonist, anti−CTLA-4, anti-PD-1 or anti-PD-L1. Group 3 consisted of TNBC patients who, after one or two prior systemic treatments for metastatic breast cancer, exhibited documented disease progression. Group 4 included ovarian cancer patients who had undergone a maximum of two prior platinum therapy courses and showed progression within 9 months following the last platinum-based regimen.

All participants needed to meet the following criteria:

1. Measurable disease as per RECIST version 1.1.
2. An Eastern Cooperative Oncology Group (ECOG)/World Health Organization performance status of 0 or 1.
3. Adequate hematological, liver, renal, and cardiovascular function.

Additionally, mandatory tumor biopsies were performed for participants in treatment groups 1 and 2, yielding two biopsies for those in Group 1 and three biopsies for those in Group 2. Patients with untreated or actively progressing central nervous system (CNS) metastases were excluded. However, individuals with a history of CNS metastasis or newly detected asymptomatic CNS metastasis were eligible to participate under specific conditions.

Participants who had previously received anti-CD137 therapy or immune checkpoint blockade therapy were eligible, provided that at least five months had elapsed since their last dose and they had no history of grade 3 or 4 immune-related adverse effects according to the NCI CTCAE criteria. Any toxicity from prior anti-cancer treatments had to be fully resolved before enrolling in the study. Furthermore, patients with known HIV infection or active viral diseases, such as hepatitis, were excluded.

#### Soluble biomarkers

Blood samples, approximately 6 ml each, were drawn from the anticubital vein at specific timepoints during the study: on Day 1, followed by weekly collections on Days 8 and 15, and subsequently every two weeks starting from Day 21. This sampling schedule was consistent throughout the initial run-in cycle, the first treatment cycle, and then every even-numbered cycle thereafter. For a detailed timeline and classification of the biomarker samples collected, please refer to Fig. 1 in the manuscript.

*Flow cytometry analysis*

Following phlebotomy at various clinical sites, whole blood samples were promptly transported under controlled ambient conditions to Covance Central Laboratories. Here, they underwent preparation and flow cytometry analysis in accordance with fit-for-purpose validated assay protocols. Data acquisition and analysis for each assay were conducted using FACSDiva software (Becton Dickinson), employing specific acquisition templates for each type of assay.

All steps of the incubation for flow cytometry were carried out at room temperature and in the dark to ensure reagent integrity. For surface staining, 50 to 100 μL of uncoagulated whole blood was used in each assay tube. This was incubated with a tailored antibody cocktail mixture for 15 minutes (refer to supplementary table 1 for a full list of antibodies). Following this incubation period, red blood cells were lysed using 3 mL of FACS Lysing Solution (Becton Dickinson) over a duration of 15 minutes. The cells were then washed with Phosphate-Buffered Saline (PBS, Sigma) to remove any residual lysis solution and cellular debris. After washing, the cells underwent fixation with 200 µL of BD Cytofix Fixation Buffer (Becton Dickinson) for 10 minutes. A subsequent wash in PBS was performed to eliminate excess fixation buffer. Finally, the cells were resuspended in 500 µL of PBS, readying them for acquisition using the FACSCanto II - 3 laser flow cytometer analyzer (Becton Dickinson).

*Statistical Evaluation of Flow Cytometry Data*

To analyze each parameter, we employed a linear mixed-effects model. In this model, individual patients were treated as random effects to account for inter-individual variability, while the timing of visits (planned visit) was incorporated as a fixed effect to assess time-dependent changes. The response variable in our model was the change from baseline for each parameter. We calculated this change using a pseudocount method:

$$\text{Change From Baseline (CFB)} = \log_2(\text{value}+1) - \log_2(\text{baseline value}+1).$$

Here, 'value' refers to the measured number of cells per microliter (cells/µL) at each time point. We set our threshold for statistical significance at a false discovery rate (FDR) of less than 0.05. This means that we considered results to be statistically significant if they had an FDR value below this cutoff.

*Cytokine analysis*

Cytokine quantification, specifically GM-CSF, IL-2, IL-6, IL-10, IL-8, IL-17A, IFNγ, and TNF-α, was conducted by Microcoat Biotechnologie GmbH, located in Bernried, Germany. This analysis employed the ELLA methodology. To assess inter-assay accuracy and precision, we utilized quality control (QC) samples at two distinct concentration levels: low-quality control (LQC) and high-quality control (HQC). These QC samples were analyzed across seventeen separate runs, which were conducted over nine days by two different analysts. The overall coefficient of variation (CV) for each cytokine at the LQC level ranged from 4.5% to 16.2%, while at the HQC level, it varied between 2.7% and 10.3%.

#### Tissue Biomarkers

Paired tumor punch needle biopsies, utilizing needles of at least 18 gauge, were collected from patients. The initial biopsy was obtained during the baseline period, ranging from Day -14 to Day -1. A subsequent biopsy was taken between Day 15 and Day 21 of the first treatment cycle. Additionally, patients in Group 2 underwent an extra biopsy within the same timeframe during the run-in cycle. For visual reference on the biomarker sample timeline, please see Fig. 1.

These biopsies were primarily extracted from easily accessible and representative tumor lesions, such as those in the liver, lymph nodes, peritoneal or abdominal cavity lesions. The process was guided by imaging techniques like ultrasound or computed tomography scans. To ensure consistency in the comparative analysis, baseline and on-treatment biopsies for each patient were, where feasible, collected from the same lesion.

Post-collection, the tumor biopsies were immediately placed into vials filled with 10% neutral buffered formalin. They were fixed in this solution for a precise duration of 24 hours, with an allowable variance of +/- 2 hours. Within 24 to 48 hours of collection, these samples were then shipped to a central pathology laboratory, HistoGenex, for paraffin embedding and subsequent analysis.

*Tissue processing and Immunohistochemistry*

For this study, ten tissue sections, each measuring 4-5 µm in thickness, were prepared from each tumor block. These sections underwent standard histological processing and immunohistochemical (IHC) procedures. Only sections containing a minimum of 50 tumor cells, as identified via hematoxylin-eosin (H&E) staining, were selected for further chromogenic assays and detailed analysis. The formalin-fixed, paraffin-embedded tissue (FFPET) sections were subjected to H&E staining using a Ventana automated system, ensuring consistent and reproducible results. Additionally, duplex chromogenic staining for CD8/Ki67 was conducted at the Good Clinical Practice Pathology Laboratory in Penzberg, employing standardized protocols for accuracy and reliability. In parallel, single chromogenic staining for the PD-L1 marker was performed at Targos using the SP263 clone, adhering to rigorous methodological standards.

*Automated image analysis*

High-resolution digital slides of the tissues were produced by scanning H&E and CD8/Ki67 stained sections using an iScan-HT scanner (Ventana Medical Systems). Subsequent manual annotations and consistency checks were performed by a pathologist, adhering to in-house guidelines that delineate criteria for normal and tumoral tissues, as well as identifying necrosis and artifacts. Necrotic regions and artifacts were excluded from analysis. The comprehensive digital slide scans underwent an automated analysis using the IRIS platform, as previously described(1). This process involved a validated, standardized algorithm specifically designed for CD8/Ki67 cell detection at the Roche Innovation Centre Munich. The analysis generated raw data encompassing quantifications and xy-coordinates for each identified object, which were then accurately stored in a standardized and validated database.

Post-analysis, metrics such as the mean count per square millimeter of tumor area (excluding significant necrotic regions and artifacts), single positive cells (CD8+Ki67-, CD8-Ki67+), and double positive cells (CD8+Ki67+) were calculated. Furthermore, the proximity of each CD8+ T cell to the nearest tumor cell was meticulously measured. All these procedures were part of an internally standardized workflow, incorporating multiple stages of quality control to ensure the reliability and accuracy of the data.

*RNA-seq analysis*

In accordance with previously established protocols(1) total RNA, inclusive of microRNA, was isolated from formalin-fixed, paraffin-embedded tissue (FFPET) utilizing the Qiagen miRNeasy FFPE kit, strictly following the manufacturer's guidelines. RNA quantification and integrity assessment were performed using the Qubit RNA Assay Kit (Thermo Fisher Scientific) and the Fragment Analyzer (Agilent), respectively. Samples exhibiting a %DV200 value below 10 were deemed unsuitable and consequently excluded from subsequent analysis.

For the construction of the sequencing library, we employed the Illumina TruSeq RNA Access methodology, starting with 100 ng of total RNA. The resulting libraries were quantified, normalized, and multiplexed in preparation for high-throughput sequencing. The Illumina HiSeq 4000 system was utilized for sequencing, adhering to a 50bp paired-end protocol and targeting a total read depth of 25 million reads per sample.

The RNA-Seq data processing involved mapping the reads to the human reference genome (hg38) using the STAR algorithm(2). Gene-level read counts were collated and normalized to account for variations in library size and composition. The analysis was restricted to genes expressed in at least three samples, with a minimum read count per million (CPM) greater than one. Seven samples exhibiting RNA degradation failed to meet our stringent quality control standards and were excluded from the analysis.

Differential gene expression analysis was conducted using the voom/limma framework (3). Our analytical model incorporated patient-specific factors and temporal variables (i.e., screening, cycle 1 day 21 for group 1, and run-in day 21 and cycle 1 day 21 for Group 2). Surrogate variable analysis(4) revealed no necessity for additional covariates in the model. To identify enriched gene signatures and pathways among the differentially expressed genes, we applied the CAMERA method, a competitive gene set test that considers inter-gene correlations(5). We defined the entire set of protein-coding genes as the background. CAMERA was then employed to analyze the ranked differentially expressed genes for each contrast, using the standard inter-gene correlation factor of 0.01.

### Supplementary Figures and Tables

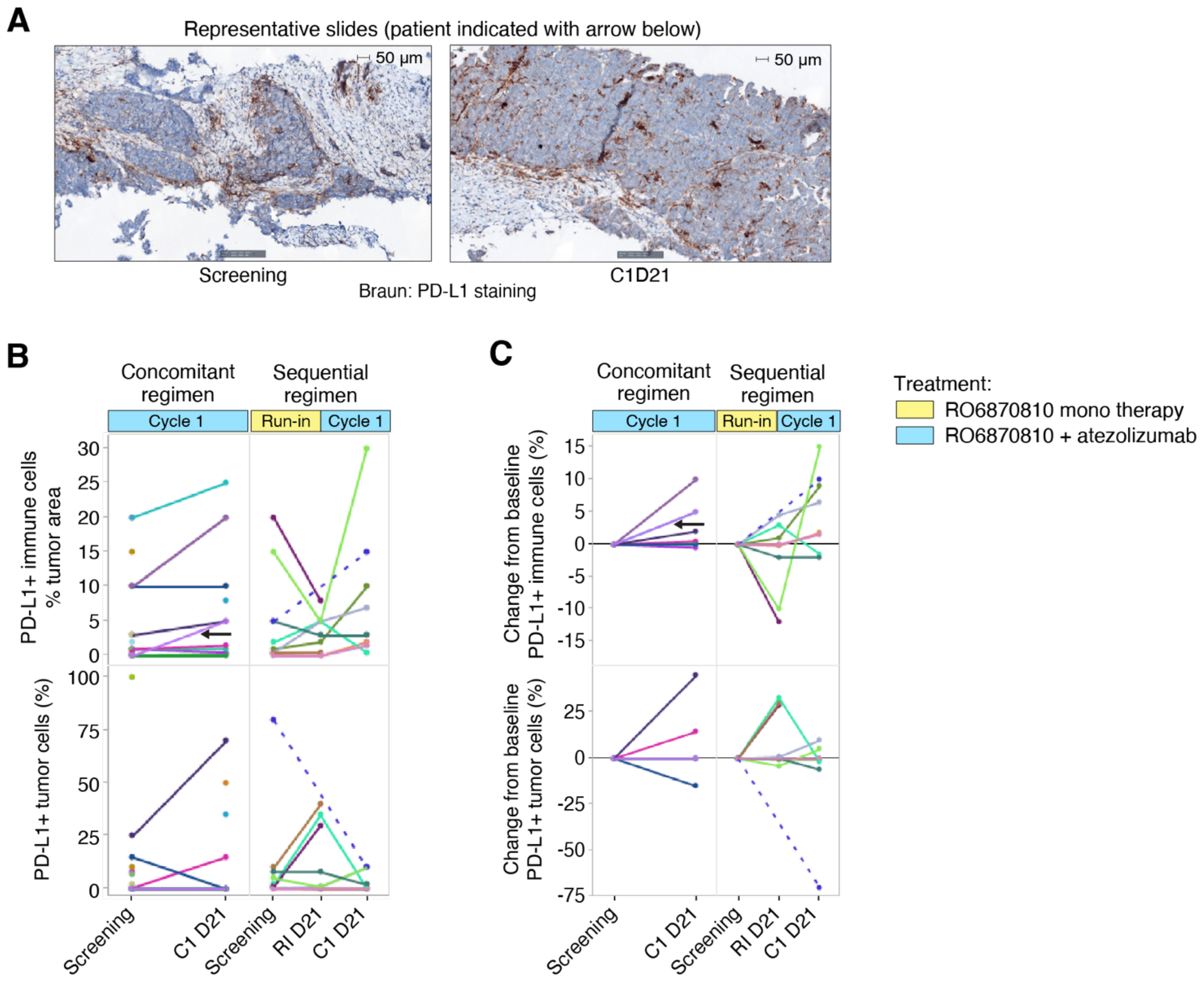


**Supplementary Figure 1: PD-L1 Expression in Tumor Tissue Characterized by IHC**

**A.** Representative digital slides of PD-L1 staining using the SP263 antibody (brown). The left panel shows the patient at baseline, while the right panel shows the same patient after one cycle of combination treatment. An increase in the density of immune cells and PD-L1+ immune cells is observed. The scale bar in the upper left corner represents 25 µm.

**B.** Percent tumor area with PD-L1+ tumor cells (top) and immune cells (bottom) for each patient at each time point. Lines connect longitudinal samples from the same patient, with different colors representing different patients. Points without lines indicate patients with missing screening or on-treatment samples. The dotted line indicates a patient with a missing run-in day 21 sample. The black arrow marks the patient whose representative digital slides are shown in Panel A.

**C.** Absolute change in the percentage of tumor area with PD-L1+ tumor cells (top) and immune cells (bottom) from the screening (pre-treatment) sample. Patients without screening or on-treatment samples are excluded. The same color coding as in Panel B is used.

**Supplementary Table 1: List of antibodies used for flow cytometry assays**

| Assay | Reagent | Manufacturer | Clone | Catalogue number |
|---|---|---|---|---|
| T, B, and NK cells | Multitest 6-color TBNK Reagent | Becton Dickinson | N/A | 337166 |
| T cell subsets | Anti-CD4 V500 | Becton Dickinson | RPA-T4 | 560768 |
| | CD3 APC-H7 | Becton Dickinson | SK7 | 560176 |
| | Anti-CD197 (CCR7) Brillant Violet 421 | BioLegend | G043H7 | 92269 |
| | Anti-CD45RA FITC | BioLegend | HI100 | 92270 |
| | Anti-CD25 PE | BioLegend | BC96 | 92274 |
| | Anti-CD8 PerCP | BioLegend | SK1 | 92271 |
| | Anti-CD127 PE-Cy7 | BioLegend | A019D5 | 92272 |
| | Anti-CD183 (CXCR3) APC | BioLegend | G025H7 | 92273 |
| Monocytes and Neutrophils | Anti-CD45 APC-H7 | Becton Dickinson | 2D1 | 560178 |
| | Anti-HLA-DR PerCP | Becton Dickinson | L243 | 7020780 |
| | Anti-CD14 PE | Becton Dickinson | MφP9 | 345785 |
| | Anti-CD16 APC | BioLegend* | 3G8 | 92188 |
| | Anti-CD11b BV421 | BioLegend* | ICRF44 | 92346 |

*Custom BioLegend Reagents
